# Analyzing Socioeconomic Factors and Health Disparity of COVID-19 Spatiotemporal Spread Patterns at Neighborhood Levels in San Diego County

**DOI:** 10.1101/2021.02.22.21251757

**Authors:** Ming-Hsiang Tsou, Jian Xu, Chii-Dean Lin, Morgan Daniels, Jessica Embury, Eunjeong Ko, Joseph Gibbons

## Abstract

This study analyzed spatiotemporal spread patterns of COVID-19 confirmed cases at the zip code level in the County of San Diego and compared them to neighborhood social and economic factors. We used correlation analysis, regression models, and geographic weighted regression to identify important factors and spatial patterns. We broke down the temporal confirmed case patterns into four stages from 1 April 2020 to 31 December 2020. The COVID-19 outbreak hotspots in San Diego County are South Bay, El Cajon, Escondido, and rural areas. The spatial patterns among different stages may represent fundamental health disparity issues in neighborhoods. We also identified important variables with strong positive or negative correlations in these categories: ethnic groups, languages, economics, and education. The highest association variables were Pop5andOlderSpanish (Spanish-speaking) in Stage 4 (0.79) and Pop25OlderLess9grade (Less than 9^th^ grade education) in Stage 4 (0.79). We also observed a clear pattern that regions with more well-educated people have negative associations with COVID-19. Additionally, our OLS regression models suggested that more affluent populations have a negative relationship with COVID-19 cases. Therefore, the COVID-19 outbreak is not only a medical disease but a social inequality and health disparity problem.

## Introduction

Since the outbreak of Coronavirus Disease 2019 (COVID-19), there are over 22.5 million positive confirmed cases and more than 375 thousand deaths in the United States (U.S.) as of 12 January 2021 (CDC 2021). With the rapid rise of new cases in Winter 2020 (November and December), timely tracking of outbreaks and their patterns at different stages as well as identifying how these differ by neighborhood characteristics are critical in addressing health disparities and providing possible strategies for mitigating the COVID-19 pandemic.

Neighborhood characteristics including race/ethnicity, elderly population sizes, socioeconomic status (SES), and geographic location contribute to health disparities in who contracts COVID-19 (confirmed cases) and their treatment outcomes (e.g., hospitalization and/or death). An effective detection and analysis of these social and economic factors and their potential impacts can help improve decision-making and health resource management plans such as the selection of COVID-19 testing sites or vaccination distribution. This study analyzed the spatiotemporal spread patterns of COVID-19 confirmed cases at the zip code level in the County of San Diego, California, U.S. and compared them to neighborhood and geographic factors from the Census Bureau’s 2018 American Community Survey (ACS). We conducted the health disparity analysis by using geographic information systems (GIS) and statistical approaches. The correlation analysis and geographic weighted regression (GWR) revealed important socioeconomic factors and spatial patterns associated with COVID-19 confirmed cases. We also analyzed the temporal factor because the outbreak patterns of COVID-19 are highly dependent on non-pharmaceutical interventions (NPIs) (social distancing, wearing masks, and stay-at-home orders), holiday events, and winter seasonal effects. This study split the San Diego COVID-19 confirmed case patterns into four stages (Figure 1) based on 7-day averages of daily confirmed cases, then compared their similarities and differences. Stage period date ranges are manually selected based on COVID-19 confirmed case growth patterns.

**Figure 1.**
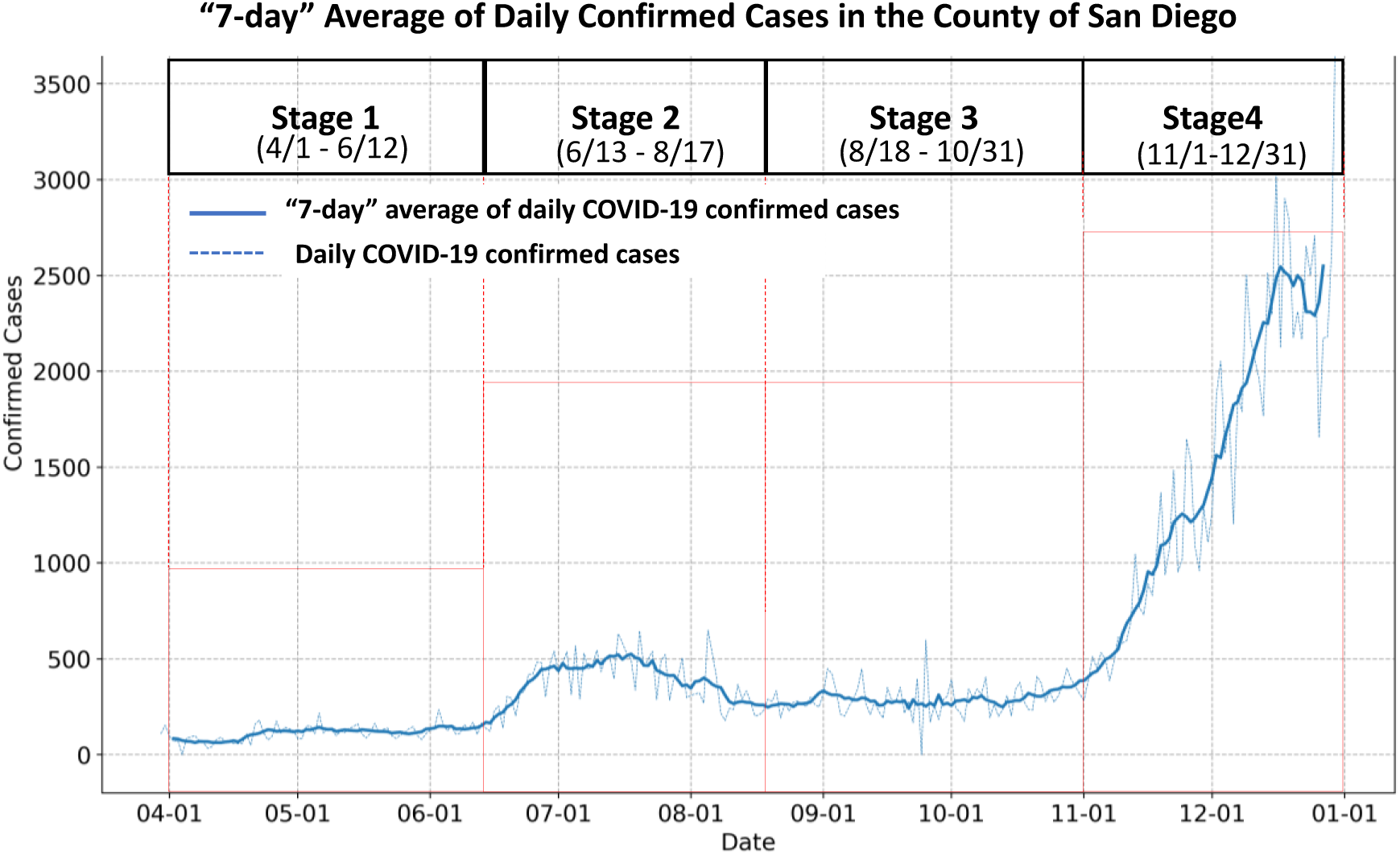
The four growth stages of 2020 COVID-19 outbreaks in the County of San Diego based on the 7-day average of daily confirmed cases.

- **Stage 1 (4/1/2020 to 6/12/2020):** Early outbreaks and slow growth rates
- **Stage 2 (6/13/2020 to 8/17/2020):** Rapid growth rates, possible association with the 6/12 reopening of gyms, bars, movie theaters, etc.
- **Stage 3 (8/18/2020 to 10/31/2020):** Reduced and stable rates, possible association with new regulations to close indoor operations at gyms, nail salons, and some other sectors
- **Stage 4 (11/1/2020 to 12/31/2020):** Rapid growth rates, possible association with the election, demonstration activities, holidays, and the cold winter season (The actual period of Stage 4 can be extended beyond 12/31/2020. We selected 12/31 as the ending date to keep similar lengths for each stage and to provide a prompt analysis result to explore possible factors connected to this fast growth period.)

To calculate the 7-day average number, we combined the daily case numbers from three days before and three days after the target date. The reason for using a 7-day average number is to remove weekly pattern effects like lower case numbers during weekend days. At each stage, we used the accumulated confirmed cases for each zip code area to compare with social and economic variables from the ACS data and identify important factors associated with COVID-19 confirmed cases at the zip code level. The detailed analysis results are illustrated in sections 2 and 3.

### Literature review: Disparities in Public Health and COVID-19

Health disparities are an important public health concern as they widen social inequality gaps in population health. Factors contributing to health disparities are complex and involve social and structural determinants, including race/ethnicity, socioeconomic status, and access to healthcare (Hooper, Nápoles, and Pérez -Stable 2020; Clark et al. 2020; Artiga et al. 2020). The current COVID-19 pandemic crisis exacerbates health disparities and inequalities resulting in negative and disproportionate negative health outcomes.

In the U.S., the effects of COVID-19 are disproportionately impacted by race/ethnicity. Disparities in COVID-19 confirmed cases and mortality are observed among African Americans, LatinX and Native Americans, and U.S. immigrants (Clark et al. 2020; Rentsch et al. 2020; Hooper, Nápoles, and Pérez -Stable 2020). As of October 2020, more than half of COVID-19 cases involved racial/ethnic minorities. Among these, 27.3% of individuals affected were Hispanics and 16.6% were African Americans while only consisting of 13% and 18% of the total population, respectively (CDC 2020). Structural inequalities associated with race/ethnicity increase the risk of COVID-19. Hispanics (21%), Blacks (17%) and American Indians/Alaskan Natives (AI/AN) (19%) are less likely than whites (13%) to see a doctor due to cost (Artiga et al. 2020). The disproportionate burden of chronic illness compounded by low socioeconomic status has widened disparities in health outcomes (Clark et all 2020). Among the non-elderly population, uninsured rates are significantly higher for Native Americans (22%), Hispanics (19%) and African Americans (12%) as compared to their white counterparts (8%) (Artiga et al. 2020). Comorbidities also tend to be higher among racial/ethnic minority groups (National Diabetes Statisics Report 2020; Kabarriti et al. 2020). For example, the proportions for comorbidities such as diabetes, chronic pulmonary disease, and kidney disease were higher among Hispanics and Blacks as compared to non-Hispanic whites (National Diabetes Statistics Report 2020; Kabarriti et al. 2020). Minorities and those with low socioeconomic status are more likely to work in industries that are open to the public during the pandemic and to live in crowded areas and multi-generational households, thereby escalating exposure to health risks and limiting options for following recommended guidelines such as social distancing and quarantines (Adamkiewicz et al. 2011; Raifman and Raifman 2020). These social and structural determinants are largely place-bound. High concentrations of racial/ethnic groups are a consequence of residential segregation (Massey and Denton 1993). Meanwhile, socioeconomic disadvantage often spatially clusters, in no small part due to racial/ethnic segregation (Sharkey 2013). Both racial/ethnic segregation and socioeconomic clustering tend to converge in specific neighborhoods, leading to resource disparities that shape access to healthcare (Kwan 2013) as well as the local availability of jobs (Wilson 2011). Consequently, the spatial concentration of disadvantage also has a direct association with poor health (Gibbons et al. 2020) including the spread of disease (Acevedo-Garcia 2001).

Currently, many health disparities and geospatial analyses of the COVID-19 pandemic focus on the global, country, or state levels while relatively few research works focus on impacts and spatiotemporal analysis at the neighborhood level (zip codes, sub-regional areas, census tracts, etc.) (Cordes & Castro 2020; Harris 2020; Lieberman-Cribbin at al. 2020). Local communities and neighborhoods play an essential role in the tracing and intervention of disease outbreaks and epidemiological research. Temporal factors also need to be considered in the spatial analysis. There are very few studies focusing on the temporal aspects of COVID-19 outbreak patterns at the neighborhood level.

In order to identify and prevent local health disparity problems associated with COVID-19 outbreaks, the State of California established the “***Blueprint for a Safer Economy***” framework on 28 August 2020. The blueprint is designed to reduce COVID-19 and specify criteria for loosening and tightening restrictions on activities in every county in California. Each county is assigned to a tier (purple: widespread, red: substantial, orange: moderate, or yellow: minimal) based on its test positivity and adjusted case rate in a weekly summary (https://covid19.ca.gov/safer-economy/). On 6 October 2020, a health equity metric was introduced in California’s *Blueprint for a Safer Economy* to determine a county’s tier. “*For a county with a population of greater than 106,000, the county must ensure that the test positivity rates in its most disadvantaged neighborhoods, referred to as the Health Equity Quartile of the* ***Healthy Places Index*** *(HPI) census tracts, do not significantly lag behind its overall county test positivity rate*”. (https://www.counties.org/csac-bulletin-article/new-health-equity-metric-released). *“The California Healthy Places Index (HPI) is developed by the Public Health Alliance of Southern California to provide overall scores on community characteristics, such as housing, education, economic, environmental, and social factors. These community conditions are called the “social determinants of health”, which inform the indicators used on the HPI platform”*. (https://healthyplacesindex.org/california-healthy-places-index-interactive-covid-19-hpi-resource-map/). The adoption of HPI in California’s “Four-Tiers” COVID-19 restriction framework indicates the importance of social and economic factors related to COVID-19 outbreaks. However, the HPI is calculated annually and therefore does not consider the dynamic temporal changes of COVID-19 outbreak patterns in local neighborhoods. Our study is one of the earliest to analyze both spatial and temporal patterns in local neighborhoods at the different stages of the COVID-19 pandemic.

### Analysis Framework and Data Processing Methods

We collected daily COVID-19 confirmed cases in this study from a public website (https://www.sandiegocounty.gov/coronavirus.html) created by the County of San Diego during the outbreak period. The county’s website provided daily updated PDF documents with COVID-19 confirmed cases at the zip code level. Our research team developed automatic PDF collection and conversion methods using Python and ArcGIS Online services to archive daily COVID-19 cases and create updated CSV files and web maps. The Python codes are available on the following Github site: https://github.com/HDMA-SDSU/ArcGIS-Python-for-COVID19-Data. The daily confirmed cases in CSV format at the zip code level and web maps are available on our COVID-19 research hub (https://hdma-sdsu.github.io/).

Figure 2 illustrates the major data collection and preprocessing procedures as well as the research framework for the social and economic factor correlation analysis of COVID-19 confirmed cases in San Diego County.

**Figure 2.**
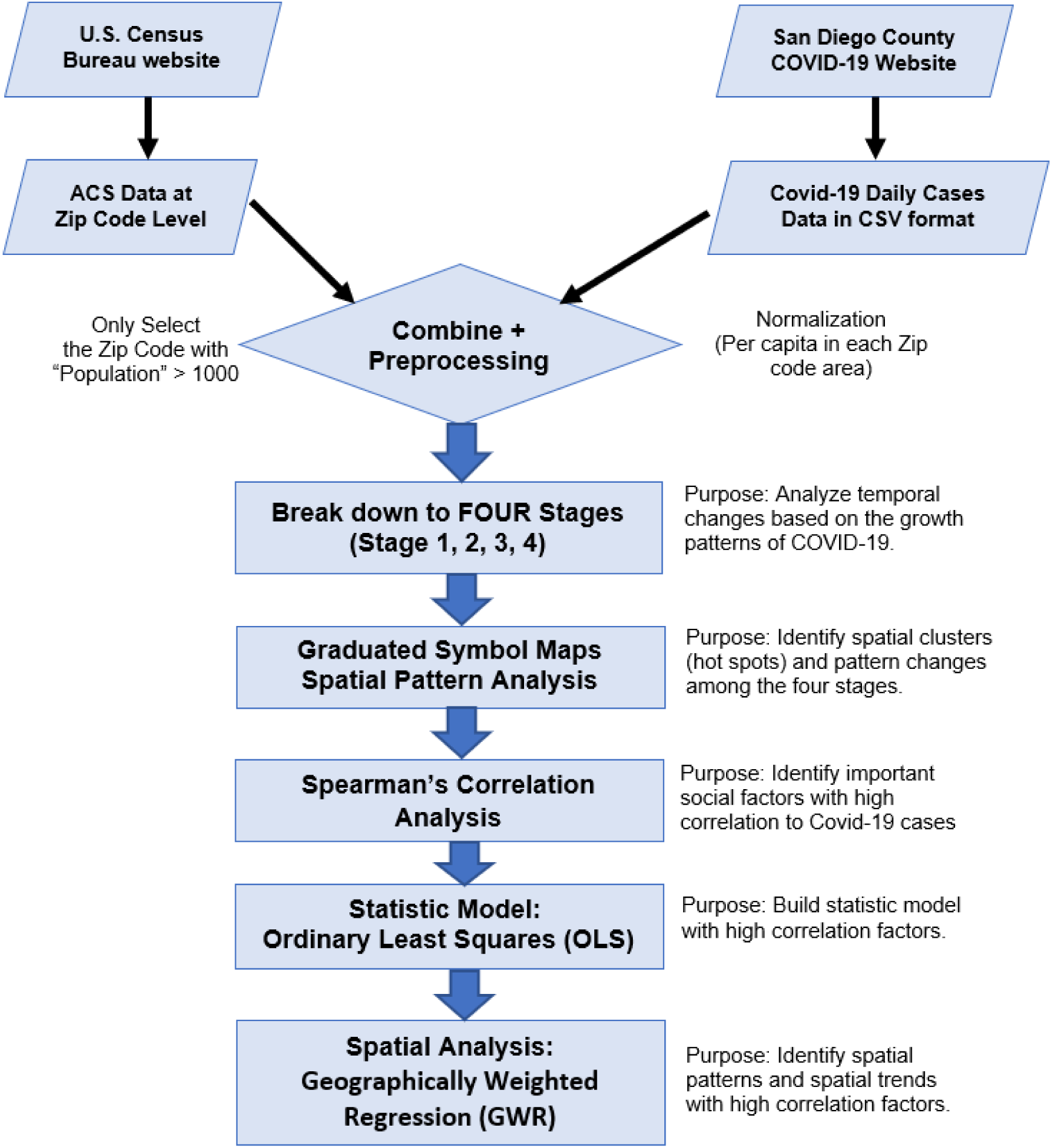
The research framework for the social and economic factor correlation analysis of COVID-19 confirmed cases in the County of San Diego.

To identify possible social and economic factors associated with COVID-19 confirmed cases in San Diego County, we utilized application programming interfaces (APIs) from the U.S. Census Bureau (https://www.census.gov/data/developers/data-sets.html) to download the American Community Survey (ACS) five-year estimates (2014-2018) data at the zip code level. We downloaded 51 socioeconomic variables including different ethnic groups, spoken languages, ages, education levels, foreign born populations, median income, total married populations, total populations with disabilities, etc. Appendix A lists all 51 ACS variables used in this study. Based on previous research, zip code areas with very small populations may not accurately reflect the characteristics of the census data. Therefore, we filtered out zip code areas with less than 1,000 residents and selected **94 zip code** areas out of **114** San Diego County zip codes. The total population of the 20 omitted zip code areas is 4,974 compared to the total population of 3,293,730 in the selected 94 zip code areas. We also normalized COVID-19 accumulated confirmed cases in each stage by the total population (2018) in each zip code area.

### Spatial Pattern Analysis (Graduated Symbol Maps) at Four Different Stages of the COVID-19 Pandemic

The first step of our analysis focused on the spatial patterns of COVID-19 outbreaks during the four different stages. We created graduated point symbol maps for each of the four stages and three additional maps showing changes between each stage. Graduated symbol mapping is one of the major cartographic representation methods used for displaying quantitative data in enumeration units such as zip code areas (Dent at all 2008). Graduated symbols are advantageous for displaying COVID-19 case numbers because they provide comparable symbol sizes between zip code areas and different stages when the same classification framework is adopted. We did not use proportional symbol methods because of difficulties in representing the small case numbers in some zip code areas.

The following maps (Figures 3 and 4) illustrate the spatial distribution patterns of COVID-19 confirmed cases (7-day average numbers) during Stages 1, 2, 3, and 4. We calculated the weekly (7-day) average numbers using the following formula:

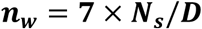

**Figure 3.**
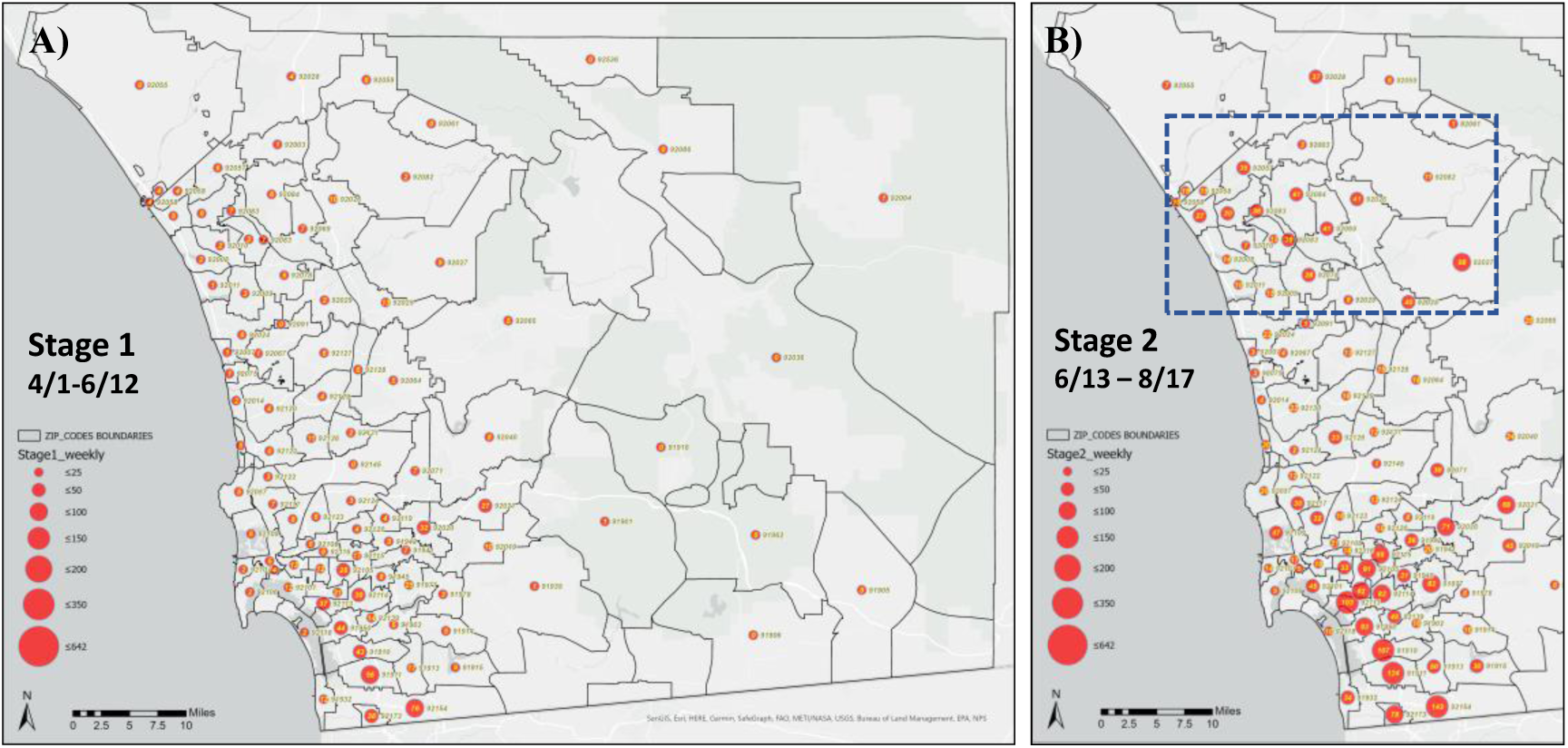
The spatial distribution of COVID-19 weekly (7-day) average confirmed cases in San Diego County in Stage 1 (A: 4/1/2020 - 6/12/2020) and Stage 2 (B: 6/13/2020 – 8/17/2020).

**Figure 4.**
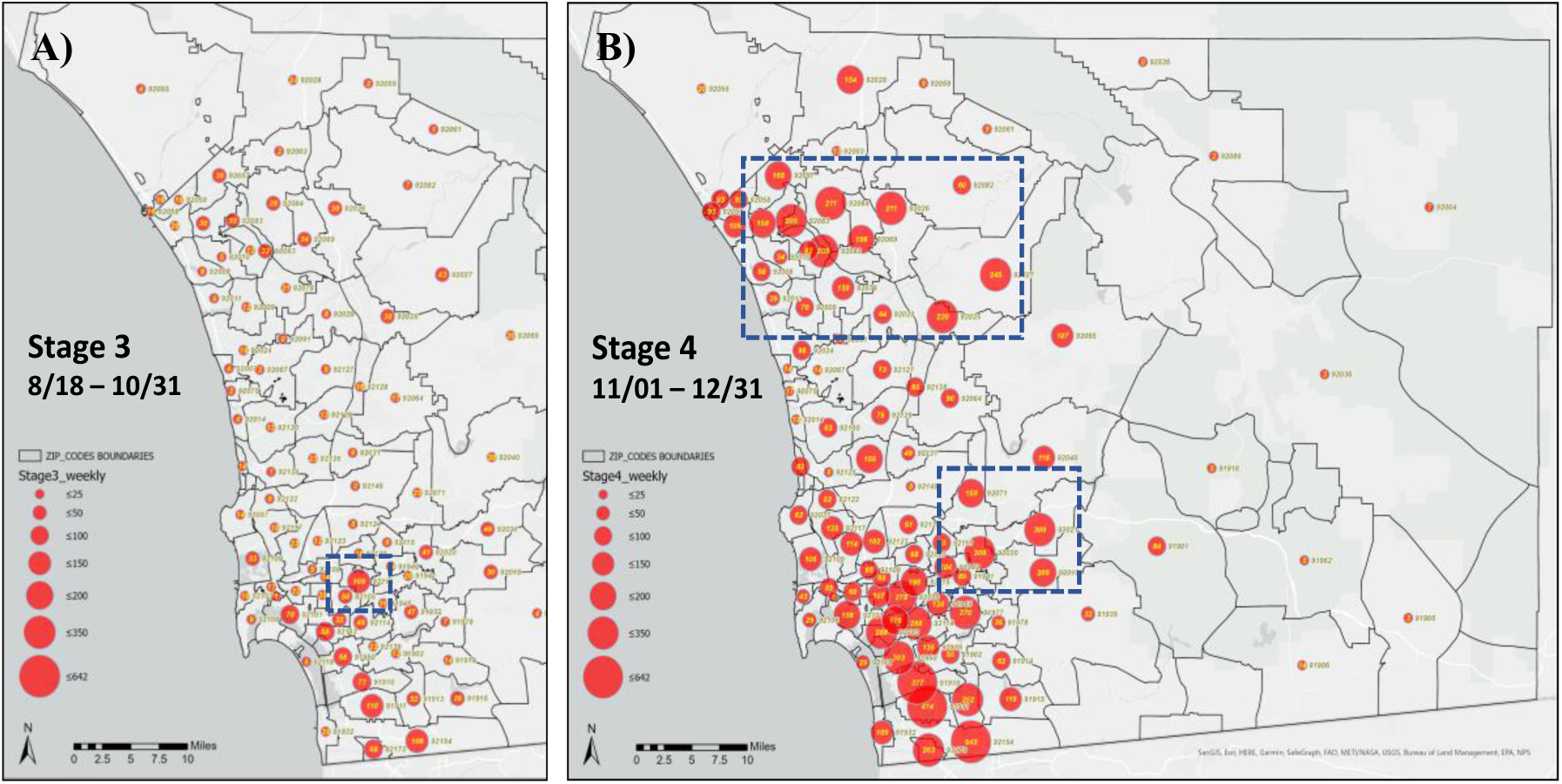
The spatial distribution of COVID-19 weekly average confirmed cases in San Diego in Stage 3 (A: 8/18/2020 – 10/31/2020) and Stage 4 (B:11/1/2020 – 12/31/2020).

Where ***n***_***w***_ represents the weekly average confirmed cases at each stage, ***N***_***s***_ is the total accumulative case numbers at each stage, ***D*** is the total days at each stage.

During Stage 1 (4/1/2020 – 6/12/2020), the majority of COVID-19 confirmed cases were located in southern San Diego County (South Bay). There were several major outbreaks in the Otay Mesa detention center (zip code 92154) in early April 2020. Stage 2 (6/13/20 – 8/17/20) shows proportional increasing case numbers in northern San Diego County areas (North County) near Escondido, Vista, and Oceanside, as well as some rural areas. The cases in southern San Diego County also increased significantly during Stage 2, especially in Logan Heights (92113), Chula Vista (91911, 91910), and Otay Mesa (92154). The growth rates of COVID-19 cases increased significantly across all zip code units in the County of San Diego. The county-wide reopening policy for gyms, bars, and movie theaters, effective 6/12/2020, offers one possible explanation for the fast growth rates in Stage 2.

In Stage 3 (8/18/2020 – 10/31/2020), the weekly growth rates slowed down in most zip code areas. One possible reason is new county regulations to close indoor operations at gyms, nail salons, and some other sectors that took effect in early August 2020. Beginning 1 September 2020, San Diego State University (SDSU) experienced an outbreak leading to increased growth rates in College Grove (92115) and downtown San Diego (92101). The SDSU outbreak started just one week after the first day of class (8/24/2020) in the Fall 2020 semester, which provided flexible student participation between online courses (around 90% of courses) and limited face-to-face instruction. The majority of the SDSU cases were among students living off-campus in the College Grove region of San Diego. In Stage 4 (11/1/2020 – 12/31/2020), all zip code areas in San Diego County had dramatically increased COVID-19 weekly average cases (bigger circles). Some areas, such as El Cajon (92021 and 92020) and Escondido (92025, 92026, 92027), had a huge increase of numbers. Similar to the Stage 2 situation, the major cluster areas are South Bay (National City, Chula Vista, Otay Mesa, and San Ysidro) and North County (Vista, Oceanside, San Marcos, and Escondido). The actual period of Stage 4 can be extended beyond 12/31/2020 as significant case growth continues into early January 2021 in San Diego County. We selected 12/31/2020 as an arbitrary ending date in order to provide prompt analysis results that explore possible factors connected to this fast growth period.

In Figure 5, we created three graduated point symbol maps to compare the changes of weekly average cases and spatial distribution patterns of COVID-19 confirmed cases between Stages 1, 2, 3, and 4. The symbol color illustrates whether the change of confirmed cases is increasing (red) or decreasing (blue). From Stage 1 to Stage 2, all zip code areas have increasing weekly average cases. North County and South Bay have the highest increasing rates. From Stage 2 to Stage 3, most zip code areas have decreasing case numbers with the exception of College Grove (SDSU), downtown San Diego, Point Loma, Solana Beach, and some rural areas. From Stage 3 to Stage 4, the changes are very similar to those seen during the Stage 1 to Stage 2 transition, where the highest increases were in North County and South Bay. El Cajon, Escondido, and South Bay have significantly increased numbers in Stage 4.

**Figure 5.**
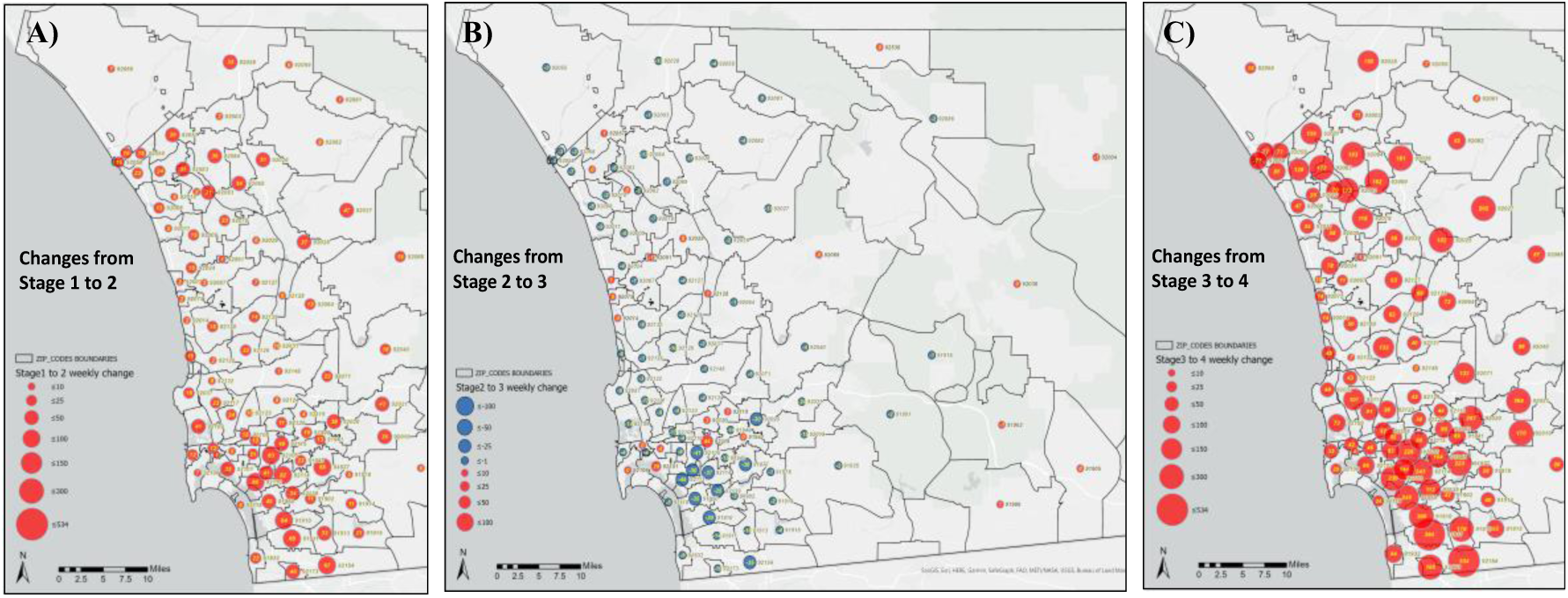
Changes in weekly average cases between Stages 1, 2, 3, and 4 in San Diego County at the zip code level (A: Stage 1 to 2; B: Stage 2 to 3, C: Stage 3 to 4) (Red: increasing cases, Blue: decreasing cases).

In general, the spatial patterns and the hot spots representative of COVID-19 outbreaks did not change too much between each stage. South Bay, North County (Escondido), and some rural areas had major clusters of COVID-19 confirmed cases. These spatial patterns may imply some fundamental health disparity issues in these neighborhoods that need to be identified and resolved to prevent future COVID-19 outbreaks. In addition to these stage change maps, our research team also created web maps showing daily increases, 7-day increases, and total accumulated cases that are published on our COVID-19 research hub. These graduated symbol maps provide an effective visualization of dynamic spread patterns of COVID-19 cases at the neighborhood level in San Diego County. The residents in each zip code area can use these maps to monitor local outbreak situations. Public health officials in the county can also utilize these maps to aid in the development of health resource management plans such as where to locate COVID-19 testing sites and distribute COVID-19 vaccines at different stages.

### Social and Economic Factor Correlation Analysis (Spearman’s Correlation)

To explore the association between COVID-19 cases and socioeconomic factors, we performed a Spearman’s correlation analysis among the 51 ACS variables, then reviewed the significance level of the Spearman’s correlation coefficient. Most social and economic factors are normalized per capita using the total population in each zip code (N=94) before implementing the correlation analysis. Table 1 illustrates 41 major ACS variables spanning six categories with significant correlation results or representative values. Results for variables with bold fonts remained consistent across the four stages. Spearman’s correlation analysis reveals important associations between COVID-19 outbreaks and socioeconomic factors at the zip code level in the County of San Diego. First, we found that **Total_population** and **Population Density** have weak associations (0.3 – 0.4) with COVID-19 cases since more populated areas or high population densities are more likely to trigger disease outbreaks in general. However, these associations are not as high as other significant social or economic factors that will be discussed later.

**Table 1.**
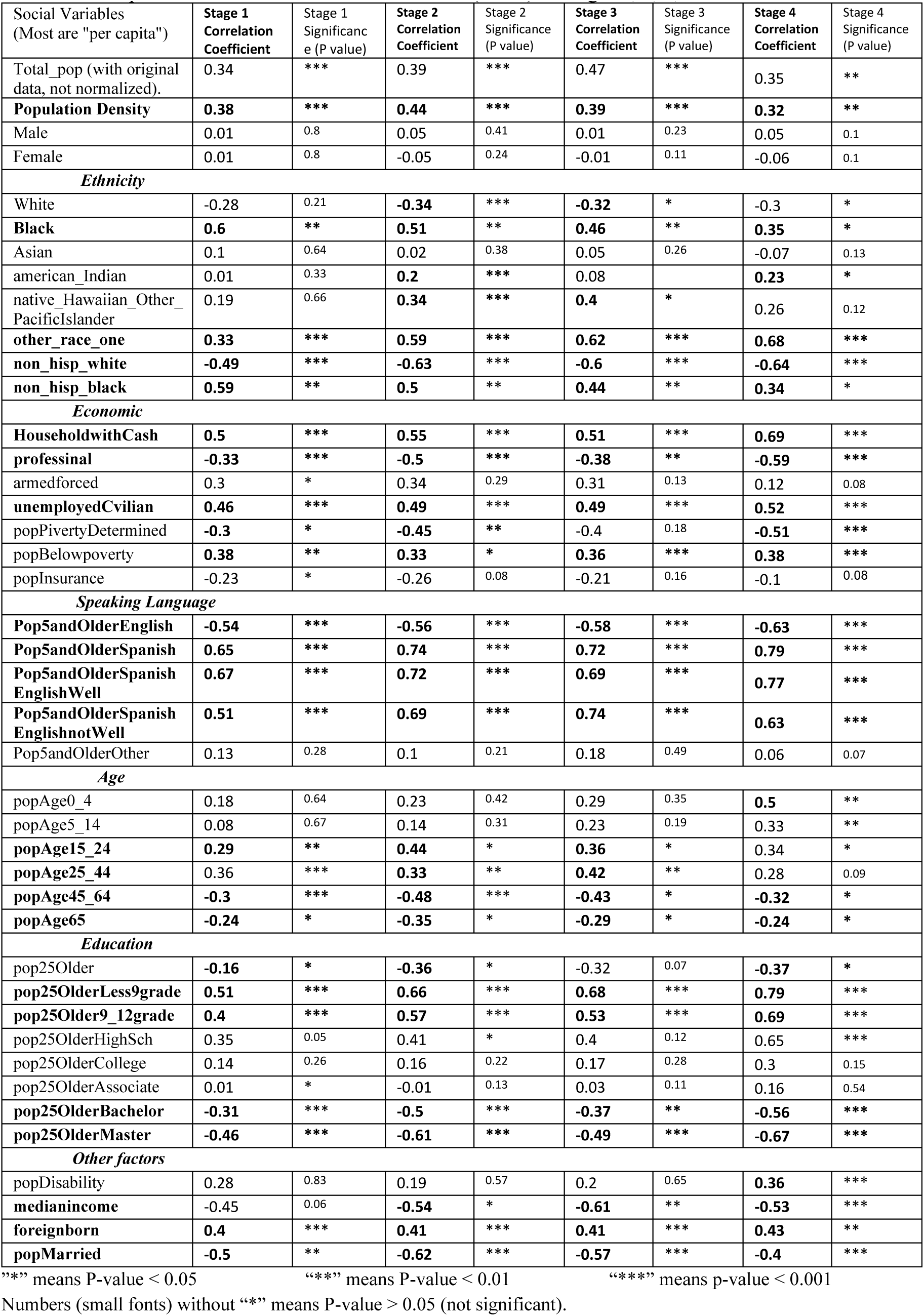
The Spearman’s correlation coefficient results (N=94) in Stages 1, 2, 3, and 4

In the ethnicity category, **Black** and **Non_Hisp_black** groups have significant positive associations (0.6 – 0.44) in Stages 1, 2, and 3. However, the association coefficient numbers in Stage 4 drop to 0.35 (black) and 0.34 (non_hisp_black). **Other_race_one** groups also have strong positive associations in Stages 2, 3, and 4 (from 0.59 to 0.68). The **Non_hisp_white** group has very strong negative associations in all four stages (from −0.49 to −0.64). Several previous studies indicate similar results for ethnic groups in New York City (Cordes, J., & Castro, M. 2020).

In the economic category, **HouseholdwithCash** (cash assistance from federal programs) and **UnemployedCvilian** have significant positive associations (0.46 – 0.69) in all stages. On the other hand, **Professional** has significant negative associations (−0.33 to −0.59) in all stages. In terms of spoken languages, **Pop5andOlderEnglish** has significant negative associations (−0.54 to −0.63) and **Pop5andOlderSpanish**, including EnglishWell and EnglishnotWell, has highly significant positive associations (0.65 to 0.79) in all stages. Regarding the age category, younger age groups (**popAge15_24** and **popAge25_44**) have weak positive associations while older and senior population groups (**popAge45_64** and **popAge65**/above) have negative associations. However, the association coefficient numbers of all population age groups decreased in Stage 4, indicating weaker associations with COVID-19 cases in November and December 2020. In terms of education level, populations with higher degree attainment (**pop25OlderBachelor** and **pop25OlderMaster**) have significant negative associations in Stages 2 and 4 (fast-growing stages). **pop25OlderLess9grade** (less than 9th grade; total population age 25 and older) and **pop25Older9-12grade** (9th through 12 grade, no diploma; total population age 25 and older) have strong positive associations in Stages 2, 3, and 4 (0.53 to 0.79). Among the other factors, **popDisability** and **foreighborn** have weak positive associations while **Medianincome** and **popMarried** have moderate negative associations.

The results from Table 1 clearly demonstrate that many important social and economic variables have strong positive or negative associations with COVID-19 confirmed cases, especially in the *ethnic group* (black, non-Hispanic white, other race), *language* (Spanish), *economic* (HouseholdwithCash, professional, and unemployed Civilian), and *education* (Less9grade/9-12grade versus Bachelor/Master degrees) categories at the neighborhood level (zip codes). We also noticed that some associations change significantly at different stages (such as Black decreasing in Stage 4, popAge0_4 increasing in Stage 4, and median-income in Stage 3) while some associations remain stable across the four stages (unemployedCvilian and popBelowpoverty). These findings warrant further examination to better understand the temporal change patterns of the variables.

Among the 51 variables, the highest association variable is Pop5andOlderSpanish in Stage 4 (0.79). In San Diego County, most families speaking Spanish are Hispanic/Latino and many have relatively large families that may live together inside a single house. Larger family sizes create a higher risk for cluster outbreaks and possible infections to seniors. Furthermore, the COVID-19 spatial cluster patterns match with the spatial distribution of Hispanic populations in San Diego County. Another very high association variable is pop25OlderLess9grade (education) (0.79). We observed a very clear pattern that regions with well-educated people (Bachelor’s and Master’s degrees) have negative associations and those with less-educated people (without Middle or High School degrees) have strong positive associations with COVID-19 cases. One possible explanation is that the first education group has a better understanding of techniques that prevent COVID-19 infection such as wearing masks, washing hands, and social distancing. Another possible explanation is that the two different education groups have very different approaches to intervention behaviors such as wearing masks and social distancing. In general, we also noticed that many variables have the highest association values at Stage 4 (a very fast-growing period) compared to the other stages, especially in the categories of languages and education. We think that the social variables in these categories are key to identifying and understanding the health disparity problems related to COVID-19 outbreaks.

Many statistical approaches, in addition to Spearman’s correlation analysis, were considered for the analysis of the data. For this study, methods were selected that best met our objective of investigating the relationships between county-wide socioeconomic explanatory variables and confirmed cases of COVID-19. In addition to our computation of the Spearman’s correlation for each set of variables, Ordinary Least Squares regression (OLS) can explain the strength of variable relationships with a linear approximation of the data. We also considered the principal component analysis (PCA) method. Ultimately, due to the nature of the data, a backward stepwise regression approach (OLS) was applied. While PCA could be of interest to reduce a large number of variables, it is not reasonably applicable for this data due to various types of potential predictors. In order to reduce the numerous predictors and multicollinearity issues while simultaneously maintaining the explanatory power of the model, the backward stepwise method was applied. Relationship strength of resulting predictors was easily explained by resultant OLS models at each stage.

### Statistical Model: Ordinary Least Squares (OLS) Regression Analysis

After conducting Spearman’s correlation analysis, the next step of the study was the creation of a statistical model to represent the dynamic spatial patterns of COVID-19 cases within San Diego County zip code areas at each of the four stages. More specifically, we sought to determine key social and economic factors within the county for each established stage. The socioeconomic data described above was analyzed using Ordinary Least Squares regression tools in R to examine the relationship between our dependent variable (COVID-19 cases) and subsets of our explanatory social and economic variables (ACS data).

Specifically, the backward stepwise regression was conducted using different nested combinations of variables to confirm that we would get the best results. In each successive model, the quality of our model was assessed using the Akaike Information Criterion (AIC) and adjusted R-squared. For every iteration, the AIC of the new nested model was compared to that of the prior to evaluate the fit. The iteration of the model that produced the lowest AIC, or prediction error, without any nonsignificant variables is the resulting model. Upon reviewing the diagnostics of this final stepwise fit, any predictors that displayed a Variable Inflation Factor (VIF) greater than ten were removed to avoid issues of multicollinearity. The backward stepwise selection process described above was then repeated to determine a final fit. This methodology preserved the explanatory power of the model, the adjusted R-squared. The process was repeated for Stages 1, 2, 3, and 4, resulting in the final OLS models shown in Table 3. Apart from Stage 2, no zip code outliers were removed in the final regression fit at each stage. The per stage final models rendered varied coefficients (Table 2).

**Table 2.** The Ordinary Least Squares (OLS) modeling variables at Stages 1, 2, 3, and 4 (Blue: positive value, Red: negative value).

| Stage 1 | Stage 2 | Stage 3 | Stage 4 |
| --- | --- | --- | --- |
| (intercept) *** | (intercept) *** | (intercept) *** | (intercept) *** |
| other_race_one *** | black . | male ** | non_hisp_white ** |
| HouseholdwithCash *** | HouseholdwithCash ** | american_Indian ** | non_hisp_black * |
| unemployedCivilian ** | professional ** | popMarried *** | popAge45_64 . |
| popBelowpoverty | popBelowpoverty ** | Total_pop | pop25OlderBachelor * |
| popInsurance *** | popMarried *** | popAge0_4 * | pop25OlderMaster *** |
| Pop5andOlderSpanish *** | Pop5andOlderSpanish *** | Pop5andOlderSpanish *** | Pop5andOlderSpanish h *** |
| pop25OlderAssociate | popAge65 ** |  | Pop5andOlderother *** |
| pop25OlderBachelor ** |  |  |  |
|  | Zip codes 92059 and 92061<br>(Pala, Pauma Valley) removed |  |  |

“.” means P-value < 0.1 “\*\*” means P-value < 0.05 “\*\*\*” means P-value < 0.01 “\*\*\*\*” means p-value < 0.001 Numbers without “\*” means not significant P-value (P > 0.1)

**Table 3.**
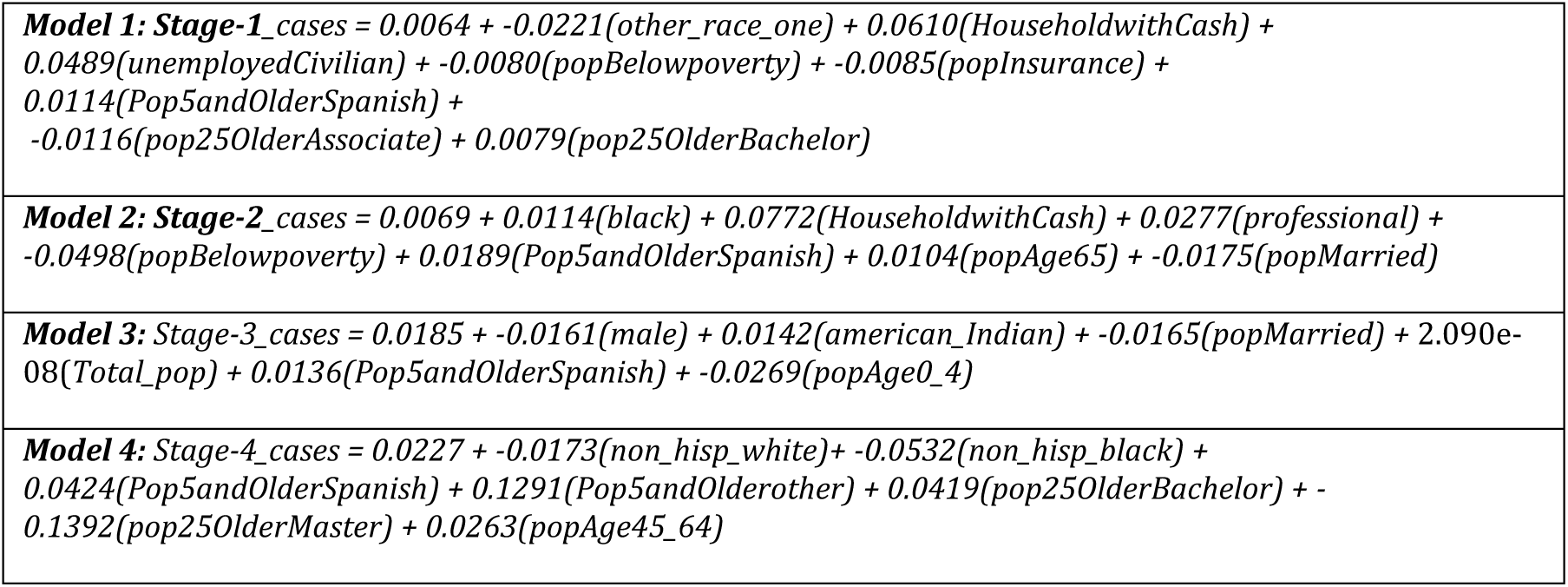
The Ordinary Least Squares regression models for Stages 1, 2, 3, and 4.

|  |
| --- |
| **Model 1: Stage-1_cases** = 0.0064 + -0.0221(other_race_one) + 0.0610(HouseholdwithCash) + 0.0489(unemployedCivilian) + -0.0080(popBelowpoverty) + -0.0085(popInsurance) + 0.0114(Pop5andOlderSpanish) + -0.0116(pop25OlderAssociate) + 0.0079(pop25OlderBachelor) |
| **Model 2: Stage-2_cases** = 0.0069 + 0.0114(black) + 0.0772(HouseholdwithCash) + 0.0277(professional) + -0.0498(popBelowpoverty) + 0.0189(Pop5andOlderSpanish) + 0.0104(popAge65) + -0.0175(popMarried) |
| **Model 3: Stage-3_cases** = 0.0185 + -0.0161(male) + 0.0142(american_Indian) + -0.0165(popMarried) + 2.090e-08(Total_pop) + 0.0136(Pop5andOlderSpanish) + -0.0269(popAge0_4) |
| **Model 4: Stage-4_cases** = 0.0227 + -0.0173(non_hisp_white) + -0.0532(non_hisp_black) + 0.0424(Pop5andOlderSpanish) + 0.1291(Pop5andOlderother) + 0.0419(pop25OlderBachelor) + -0.1392(pop25OlderMaster) + 0.0263(popAge45_64) |

In Stage 2, the studentized residuals and Q.Q. plot of the initial backward stepwise result suggested some outliers. Zip codes 92059 and 92061 were deemed appropriate to eliminate. Zip code 92059 contains Pala, the largest reservation in the county, and zip code 92061 is the adjacent Pauma Valley, considered part of the Pala region. Both zip codes contain casinos. According to U.S. Census Bureau, the population of 92059 is 47.6% American Indian/Alaskan Native (Non-Hispanic). Using data from the San Diego County COVID-19 website, analysis of San Diego County cumulative cases by ethnicity revealed that this group had the lowest growth rate in cases as a percentage of their population. Tribes are separate sovereign governments that self-regulate their local economies. Moreover, tribal governments are not subject to the state’s COVID-19 restrictions and reporting protocols and may subsequently defy broader county-wide trends. The presence of casinos in these zip codes were anomalous given their exemption from business closures required by local health orders. The casinos have been open in some capacity since May 2020 and limited COVID-19 information has been provided by the tribal governments. Anomalies also appeared in these zip codes in the other stages but were not significant and did not warrant removal from the models.

Upon comparison of the OLS results for the four stages, each stage was found to have a unique combination of independent variables, which may indicate significant distinctions between outbreaks in different stages (Tables 2 and 3). **Pop5andOlderSpanish** (Spanish-speaking population) was the only variable identified by all four stages. For each stage, the p-value for the parameter **Pop5andOlderSpanish** was less than 0.05, thereby explaining a significant amount of the variation within the model and suggesting a strong association with the linear approximation of our data. The estimate for each stage also showed a positive relationship between the number of cases and the predictor variable. This finding affirmed published county trends of graver COVID-19 impacts on minority communities. The surrogate predictor, **Pop5andOlderSpanish**, highlights the disproportionate effect of the virus on the LatinX community during every stage.

In contrast, the **non_hisp_white** predictor had a significant negative parameter estimate in Stage 4. This follows with the steady increase of the correlation coefficient for this predictor across the four stages as seen in Table 1. When all other predictors in the Stage 4 model remain constant, a population increase of one person of **non_hisp_white** identity decreases the COVID-19 case count by a negative factor between 0.49 (Stage 1) and 0.64 (Stage 4). The inverse relationship between COVID-19 cases and the proportion of whites reinforces observations that the adverse effects of the pandemic are greater in communities with higher proportions of minorities. Larger white populations suggest less prevalence of cases while cultural indicators such as **Pop5andOlderSpanish** and **Pop5andOlderother** remained positively significant to the model. The presence of these predictors versus specific *race/ethnicity* variables suggests that the *language* categories may be more indicative of cultural variance.

In Stages 1 and 2, **popBelowpoverty** and **householdwithcash** both prove significant to our model, but, interestingly enough, the variables had opposite relationships. The **popBelowpoverty** predictor had a negative association, meaning that a unit increase in the total population below the poverty level would decrease the model’s case prediction in that stage, assuming all other predictors were held constant. Conversely, the **householdwithcash** predictor displayed a positive association, indicating that the number of predicted cases increases with each unit added to the number of households that received cash assistance. In Table 1, the **popBelowpoverty** variable was shown to have a positive Spearman’s correlation coefficient in all stages, so its negative estimates in Stage 1 and 2 of the model may stem from multicollinearity, although VIF was tested for all predictors and found to be a non-issue. Regardless, these socioeconomic status variables have a definite impact in the model’s beginning stages eventually disappear in Stages 3 and 4.

Other predictors, specifically *education* category variables, were also significant to our models of Stages 1 and 4. **pop25OlderMaster** proved to have a significant negative relationship within our Stage 4 model and **pop25andOlderBachelor** proved positive in both stages. Curiously, the **pop25OlderAssociate** variable was also negative in Stage 1. Once again, there may be some issues of multicollinearity in this stage but none of the model’s rules were violated. Nevertheless, it is clear that educational attainment is highly important to San Diego County COVID-19 trends. The Stage 4 model predicted that an increase in higher education levels, above a Bachelor’s degree, will decrease the number of cases when all other variables are held constant. Higher education levels are commonly associated with higher salaries and greater wealth, suggesting that more affluent populations have a negative relationship with COVID-19 cases and thus a lower risk than less affluent communities.

### Geographically Weighted Regression (GWR) Analysis

One disadvantage of OLS regression is that the OLS model assumes that values of the correlation coefficients are distributed randomly across the study area. However, we believed that the ACS socioeconomic variables may display strong spatial variation at the neighborhood level. In other words, the strength of each variable’s effects might vary across the study area. Consequently, we leveraged Geographically Weighted Regression (GWR) to produce more informative results regarding the variation of parameters across the study area. Compared to the spatial stability of the OLS model, the variables in the GWR show spatial non-stationarity.

After applying correlation and OLS regression to socioeconomic factors and COVID-19 cases, we identified variables that are highly correlated with COVID-19 and built a multilinear model using those variables. To unlock the spatial variation in the associations of the selected socioeconomic factors to COVID-19, we employed GWR along with Spatial Autocorrelation. Commonly viewed as an exploratory tool (O’Sullivan and Unwin 2010), GWR allowed us to generate empirically determined coefficients for each observation, producing a more nuanced assessment of the association between our dependent variable (COVID-19 cases) and independent variables (selected social factors) at a local level (Becker 2019; Fotheringham, Brunsdon, and Charlton 2003).

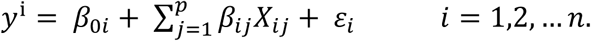

Where i represents the zip code, *y*^*i*^ is the COVID-19 case prediction, *β*_0*i*_ is the intercept, *ε*_*i*_ is the random error, *β*_*ij*_ is the regression parameter and *X*_*ij*_ is the value of the explanatory parameter. Based on such analysis, we built a local model of the impact of COVID-19 over the County of San Diego.

#### Step 1: Spatial Autocorrelation

In this study, we applied spatial autocorrelation to the number of cumulative COVID-19 cases in the different stages using the Moran’s I method. The results are shown in Table 4 below. Based on the Moran’s I analysis, we found that Z-scores in all four stages are very high with significant p-values. The COVID-19 data in all stages showed clustered distributions and strong spatial autocorrelation. The Moran’s Index from Stage 1 to Stage 4 are all above 0.3. For reference, higher index values signify more concentrated spatial distributions and more significant correlations. The result is consistent with the actual situation; the numbers of COVID-19 cases are more correlated and severe in clustered areas. Next, we performed GWR to explore the spatial regression relationship between COVID-19 cases and selected socioeconomic variables.

**Table 4.** Moran’s Index results for COVID-19 cases in different stages:

| Stages | Moran's I | Z-score | P-value | Distribution |
| --- | --- | --- | --- | --- |
| Stage 1 | 0.5114 | 8.4250 | *** | Clustered |
| Stage 2 | 0.3199 | 5.5683 | *** | Clustered |
| Stage 3 | 0.3784 | 6.1946 | *** | Clustered |
| Stage 4 | 0.4285 | 7.2348 | *** | Clustered |

#### Step 2: GWR analysis and explanations

To explore the local potential spatial variation, the same independent variables in the OLS model (Table 2) were used to build the GWR model. We tested the GWR model in each stage. Using these socioeconomic variables, GWR explained nearly three-quarters of the variation in our dependent variable in Stages 1, 2, and 4. For instance, the adjusted R-squared value increased slightly from 0.74 in the OLS to 0.76 in the GWR model in Stage 1. The AIC value of -755 was substantially low, demonstrating the robustness of the GWR model. The adjusted R-squared significantly increased from 0.45 to 0.65 in Stage 2. The adjusted R-squared then decreased at Stage 3, possibly due to heightened control of the government and stability of the COVID-19 pandemic. The adjusted R-squared is 0.79 in the GWR at Stage 4, which increased from 0.73 in the OLS model. Special events like the Presidential election may account for the surge of COVID-19 cases when infective cases show strong spatial patterns and high correlations with those social factors again at Stage 4. The goodness of fit for OLS and GWR in each stage is displayed in Table 5 below.

**Table 5.** The measures of Adjusted R^2^ for OLS, GWR models for COVID-19 cases in San Diego County:

| Model | Stage 1 | Stage 2 | Stage 3 | Stage 4 |
| --- | --- | --- | --- | --- |
| OLS | 0.72 | 0.45 | 0.48 | 0.73 |
| GWR | 0.74 | 0.65 | 0.55 | 0.79 |

Figure 6 illustrates the local GWR coefficients for the variable “Pop5andOlderSpanish” (Spanish speaking population) that is identified at each stage. Thus, this factor is critical in explaining the distribution of COVID-19 in the County of San Diego. The coefficient of the Spanish-speaking population is high in the southern region in the GWR model for each stage, which means the Spanish-speaking group is an influential factor in explaining COVID-19 case distribution in the southern area of San Diego County, particularly in Chula Vista and Otay Mesa. Figure 7 illustrates the spatial distribution of local R-squared for each stage in San Diego County. Overall, zip codes in the southwest parts (Chula Vista, Downtown San Diego) of San Diego County had relatively high local R-squared, indicating good predictions by the model in those areas. By contrast, the local R-squared values were relatively low in the northwest areas in San Diego County, which suggests poor performance of the GWR model across these regions. The GWR model provided good explanation rates for COVID-19 case distribution in different zip codes at Stage 1. In the second stage, the GWR model shows a higher explanation rate in the southern part of San Diego County as compared to the northern areas. In the third stage, the GWR model has a poor fit throughout the entire San Diego County due to heightened government control and reduced COVID-19 spread. At Stage 4, the GWR model has an improved fit throughout the County of San Diego as compared to the other stages. The explanation rate of the model increases from north to south and keeps the highest rates in the southern areas of Chula Vista and Otay Mesa. The pattern is consistent with the distribution of COVID-19 cases. That is, the GWR model fits better in the regions with more infective cases. When building the GWR model, close attention was paid to collinearity issues in explanatory variables, which can cause the failure of the GWR model. Moreover, the OLS model is a prerequisite step to building a reasonable GWR model.

**Figure 6.**
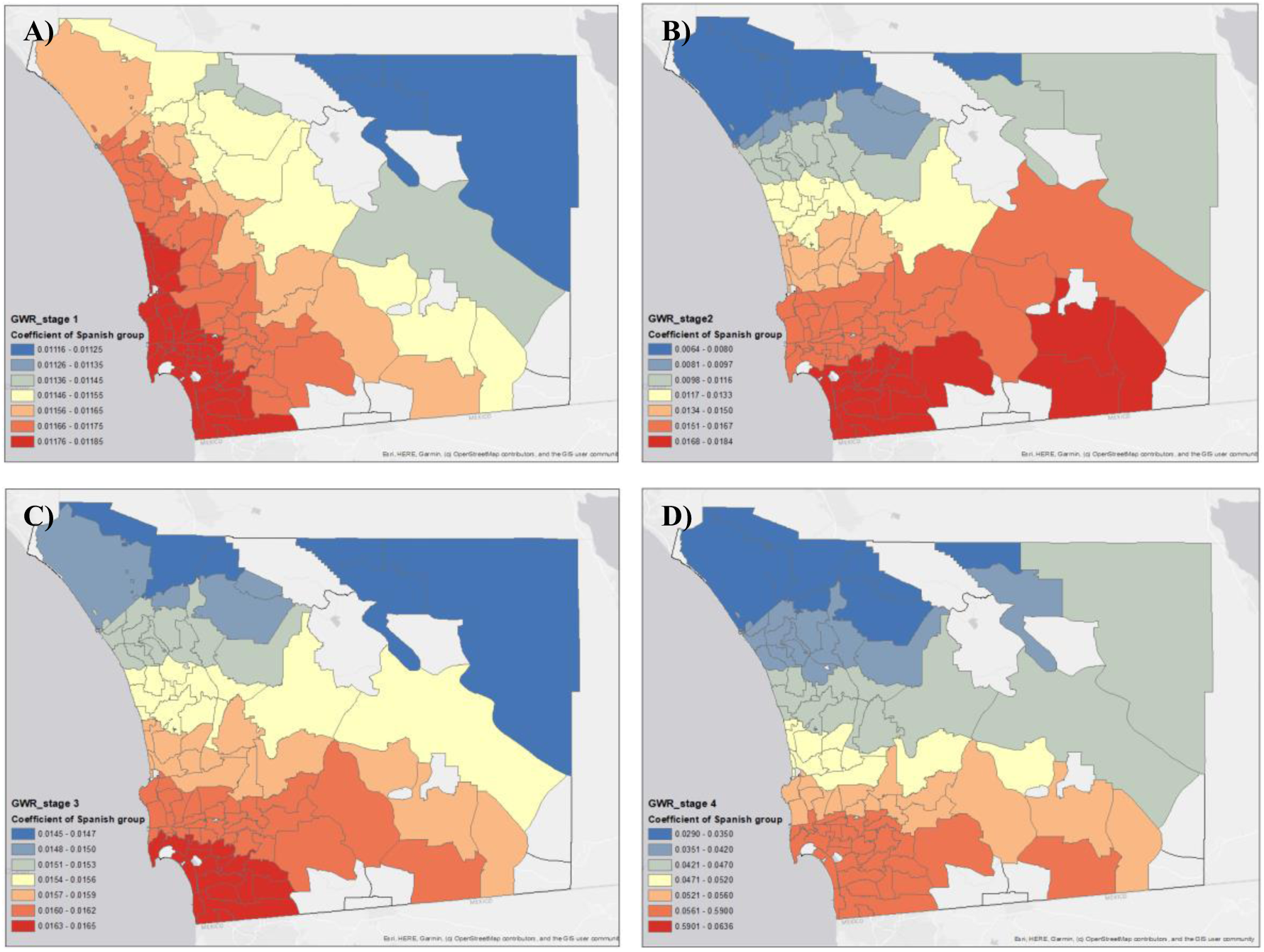
The coefficients of “**Pop5andOlderSpanish**” (Spanish-speaking) in the GWR model for COVID-19 at different stages: A) Stage 1 (4/1/2020 – 6/12/2020), B) Stage 2 (6/13/2020 – 8/17/2020), C) Stage 3 (8/18/2020-10/31/2020), D) Stage 4 (11/1/2020 – 12/31/2020).

**Figure 7.**
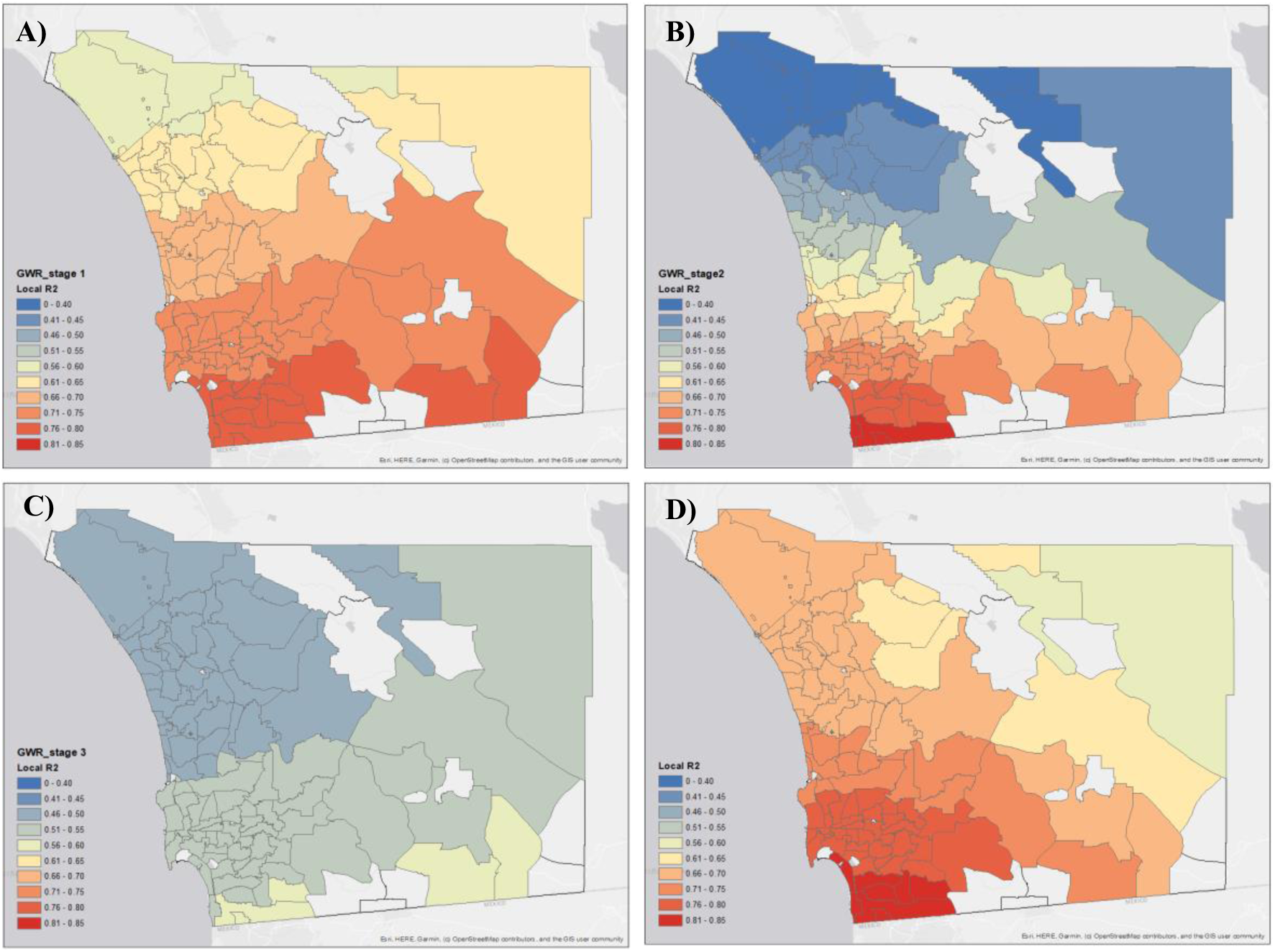
The GWR model local R-squared (R^2^) for different stages: A) Stage 1 (4/1/2020 – 6/12/2020), B) Stage 2 (6/13/2020 – 8/17/2020), C) Stage 3 (8/18/2020 - 10/31/2020), Stage 4 (11/1/2020 – 12/31/2020). Empty cells indicate areas with low population, which were not considered in this research.

From a spatial perspective, the general GWR trends among the four stages are very consistent. South Bay areas in southern San Diego County have the highest spatial clustering effects and the strongest association between COVID-19 cases and identified social and economic variables. From a temporal perspective, the adjusted R-squared value in Stages 1, 2, and 4 are much higher than in Stage 3. The “South Bay Cluster” had very strong effects during the initial stage and the fast growth stages of the pandemic. Stage 3, characterized by decreasing COVID-19 case rates, had the lowest adjusted R-squared value which indicates that the slowdown in the spread of the COVID-19 weakened the spatial clustering effects. Stage 4, identified by the second wave of rapid COVID-19 case growth, had relatively high adjusted R-squared values compared to Stages 2 and 3, meaning that COVID-19 displays a strong spatial pattern and high correlation with social factors when cases surge.

## Discussion

This study provides a comprehensive spatial and temporal analysis framework for the study of COVID-19 outbreaks at the neighborhood level (zip codes). Although the focus of this research was limited to COVID-19 confirmed cases in the County of San Diego, the temporal patterns (four stages) identified in San Diego County are similar to those in other Southern California counties (Orange County, Riverside County, LA County, etc.), based upon this CovidPulse visualization: https://www.esri.com/arcgis-blog/products/arcgis-living-atlas/mapping/covidpulse-update-grid-view/. Our analysis framework could easily be applied to many counties in Southern California if daily COVID-19 cases are available at the zip code level. Since April 2020, our research team has met weekly with public health staff in the County of San Diego to communicate and share our findings and suggestions. Additionally, we received feedback and suggestions from the public health staff regarding health disparity issues. The spatiotemporal analysis and social variable analysis frameworks can help local health agencies provide more effective health resource management and communication strategies related to COVID-19 outbreaks such as setting up more testing sites, targeting high risk audiences, and planning vaccination sites.

One important clarification for this study is that “*correlation does not mean causation*”. Although we have identified several important social and economic variables with very strong positive or negative correlation values (0.74 - 0.79), readers should not interpret that these variables are the direct reasons causing COVID-19 outbreaks. For example, the spoken language variable (Spanish-speaking) is not the reason behind COVID-19 outbreaks. However, this variable is closely related to larger family sizes common among Hispanic populations, which may be the key factor triggering more COVID-19 cases in these neighborhoods.

Another important issue is how to predict the spatiotemporal change patterns of COVID-19 outbreaks. When comparing the four stages of the study, there are many possible events or factors impacting the growth patterns of COVID-19 cases including local policy changes and orders, non-pharmaceutical interventions (NPIs), social events, family gatherings during holidays, and winter weather conditions resulting in more closed windows and less ventilation. Furthermore, different neighborhoods have different responses and reactions to these factors and other intervention orders. Most traditional disease prediction models, like SEIR (Yang et al. 2020) and GLEAM (http://www.gleamviz.org/), do not consider spatial variations and spatial health disparity issues in local communities. Therefore, we must develop a new spatially oriented disease spread model to consider major social and economic variables at different neighborhood levels and different stages. The new model should have a dual focus on the identification of hotspot regions and the impact to vulnerable populations from a spatiotemporal perspective.

Data privacy is also a major challenge for COVID-19 outbreak analysis. Although local government agencies have detailed resident addresses for each confirmed or hospitalized COVID-19 case, only aggregated data can be released to the public or researchers in order to protect patient privacy. San Diego County is one of only a few counties in U.S. willing to provide zip code level aggregation data. In our public web maps, we must also suppress the data if the case number in a zip code area is less than five.

## Conclusion

This study illustrates the important role of geography in understanding the spread of COVID-19 in the County of San Diego. The spatial clustering patterns of COVID-19 hotspots can be correlated with unique social and economic characteristics (Spanish-speaking, less education, low income, etc.). Current public health orders or NPI methods issued by state or local government agencies are typically designed for the whole county or the whole state rather than customized for high-risk local communities. To provide more effective public health intervention methods for COVID-19, we need to create geographically targeted strategies for non-pharmaceutical interventions and the resolution of fundamental healthy disparity problems in these neighborhoods.

Both state and local governments need to adjust their public health orders and interventions with consideration to the actual needs of vulnerable local communities, perhaps by providing more financial supports and incentives for large families, clear quarantine procedures and safety guidance for Hispanic/Latino populations, and free high-speed Internet services for quarantined families. We have to implement effective social and economic approaches to solve COVID-19 outbreak problems.

There are still several hidden challenges to providing effective non-pharmaceutical interventions (NPIs), like social distancing, mask wearing, and stay-at-home orders at the local neighborhood level. The first hidden challenge is the stigma of COVID-19. Evidence suggests that COVID-19 causes significant psychological distress in the forms of anxiety, depression, and fear of uncertainty among individuals affected by COVID-19 as well as their family members and healthcare workers (Duan, Bu, and Chen 2020; Guo et al, 2020; Peprah and Gyasi 2020). Stigma of having COVID-19 is a major source of distress and a key challenge that must be overcome to implement an effective public health response. Stigmatization can negatively impact motivation and behaviors of individuals affected by COVID-19 and may result in hiding or underreporting symptoms, failure to report important medical history (Peprah and Gyasi 2020) or seek a COVID-19 test (Earnshaw et al. 2020). Immigrants with limited access to healthcare and fear of potential legal repercussions have higher risk of becoming infected and developing severe COVID-19 symptoms (Clark et al. 2020). Another hidden challenge is the lack of proper COVID-19 prevention knowledge associated with certain race/ethnicity, age, gender, and socioeconomic status groups. African Americans and Hispanics, people age 55 and younger, men, and people with lower incomes had less knowledge about COVID-19 (e.g., symptoms, how it is spread) as compared to their counterparts (Alsan et al. 2019). Lack of knowledge about COVID-19 increases the risk of non-health seeking behaviors or failure to adopt recommended health behaviors.

Hence, there has been an increasing need to implement stigma-mitigation strategies and interventions. First, community engagement strategies might be effective in facilitating community residents’ participation in COVID-19 programs and enhancing social cohesion to combat stigma (Duan, Bu, and Chen 2020; Logie and Turan 2020). Second, as lack of knowledge or misinformation is often the cause of fear and stigma, implementing educational efforts to increase knowledge is important. Media can be an effective method to reduce stigma. Providing up-to-date knowledge and debunking myths and misperceptions related to COVID-19 via use of media interventions can reduce stigma as well as promote health seeking behaviors such as COVID-19 testing in the public (Duan, Bu, and Chen 2020; Earnshaw et al. 2020). Such messages need to be provided in a culturally appropriate manner, with consideration to the languages and characteristics of stigmatized groups or communities (Peprah and Gyasi 2020).

Understanding the spatiotemporal patterns of the COVID-19 pandemic is a grand challenge for geographers and geospatial data scientists. There are many hidden mechanisms and correlations to be discovered and analyzed. This study illustrated some potential research directions. However, we need to integrate more data from different resources and create an information synthesis approach (Sui, 2016). For example, combing the human mobility data from Apple Mobility Reports, Google Community Mobility Trends, and SafeGraph mobility data might help us to analyze the relationship between COVID-19 outbreaks and travel patterns between counties or local areas. By using social media data such as Twitter and Facebook comments, we can analyze public opinions and behaviors related to government orders or policy changes.

To enable a data synthesis research framework for a COVID-19 outbreak study, geographers need to collaborate with many experts including data scientists, epidemiologists, public health officials, health practitioners, sociologists, statisticians, social workers, and other domain experts. The COVID-19 outbreak is not only a medical disease and public health crisis but also a social inequality and health disparity problem. We must build transdisciplinary research teams in order to develop effective solutions for mitigating and resolving COVID-19 outbreaks.

## Data Availability

We collected daily COVID-19 confirmed cases in this study from a public website (https://www.sandiegocounty.gov/coronavirus.html) created by the County of San Diego during the outbreak period. The county's website provided daily updated PDF documents with COVID-19 confirmed cases at the zip code level. Our research team developed automatic PDF collection and conversion methods using Python and ArcGIS Online services to archive daily COVID-19 cases and create updated CSV files and web maps. The Python codes are available on the following Github site: https://github.com/HDMA-SDSU/ArcGIS-Python-for-COVID19-Data. The daily confirmed cases in CSV format at the zip code level and web maps are available on our COVID-19 research hub (https://hdma-sdsu.github.io/). The demographic data used in this research is from the U.S. Census Bureau (https://www.census.gov/data/developers/data-sets.html).

## Acknowledgement

Thanks to the creation of the public COVID-19 data sharing website and review suggestions by the staff and epidemiologists in the County of San Diego, Health and Human Services Agency, Public Health Services, Epidemiology and Immunization Services Branch and Community Health Statistics Unit.

## Appendix A

The full list of 51 American Community Survey (ACS) variables used in this study: five-year estimates from 2014 – 2018 at the zip code level.

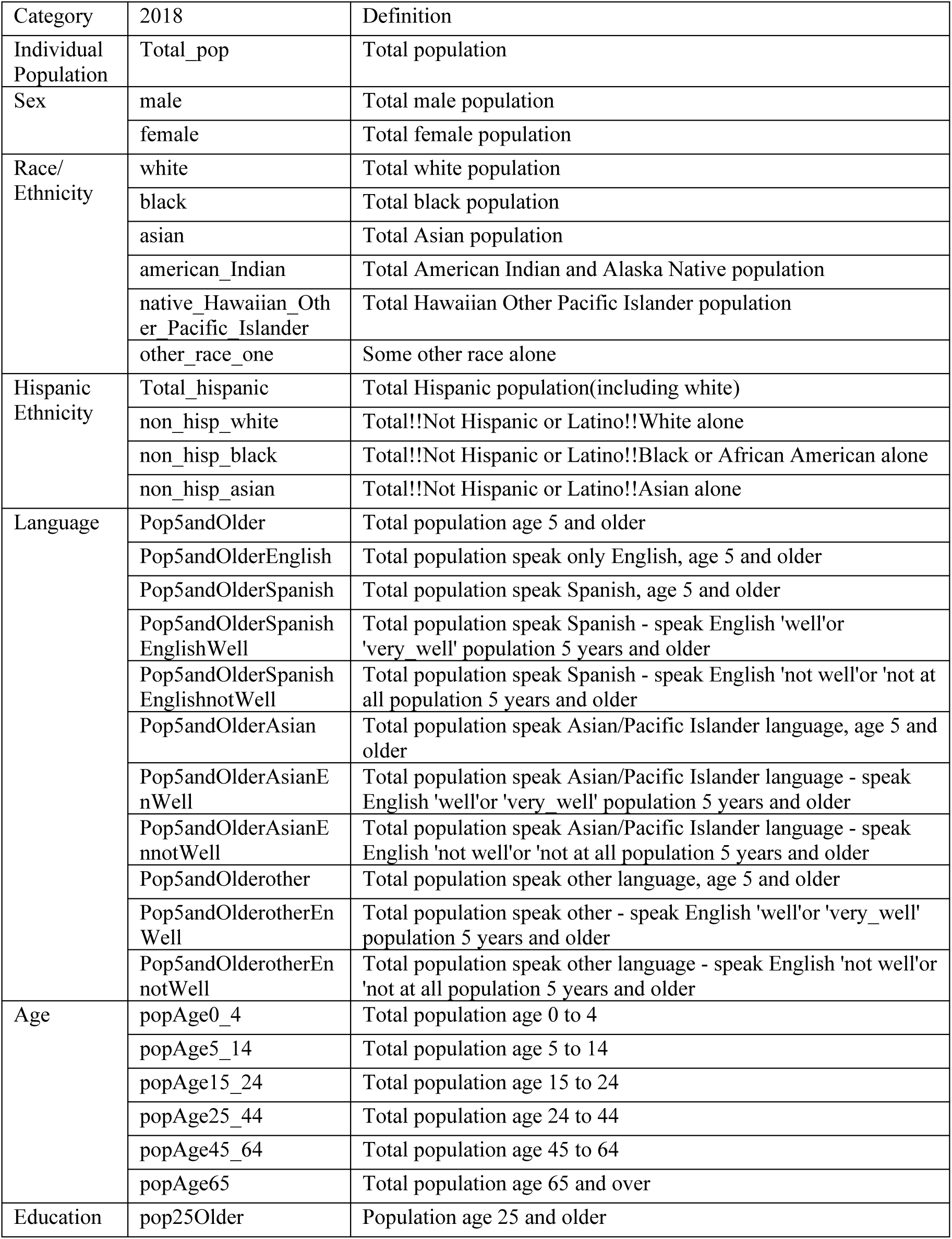
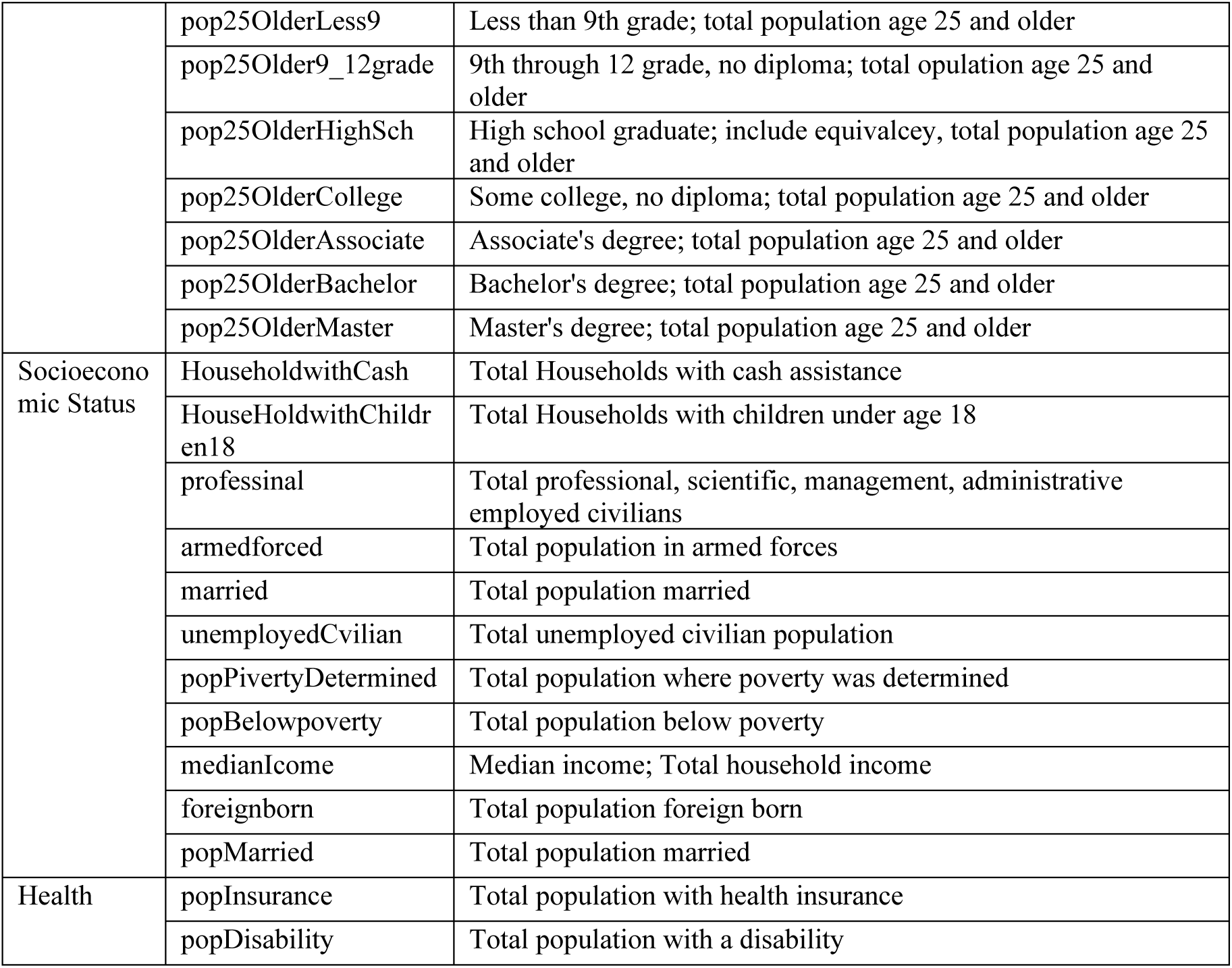

